# Using Simulated Encounters with Standardized Patients to Teach Medical Students to Address Implicit Biases and Microaggressions in the Clinical Setting

**DOI:** 10.1101/2024.06.01.24308315

**Authors:** Rachel Thommen, Harli Weber, Katherine Lo, Deelan Ayhan, John Vellek, Lior Levy, Sarah Smith, Christopher Hoke, Redab Alnifaidy, Danielle Vargas, Katharine Yamulla, Penny Liberatos, Mill Etienne, Pamela Ludmer

**Affiliations:** School of Medicine, New York Medical College, Valhalla, NY USA; School of Health Sciences and Practice, New York Medical College, Valhalla, NY USA

**Keywords:** Implicit Bias, unconscious bias, medical education, experiential learning

## Abstract

**Purpose:** Structural obstacles in healthcare related to social, economic, and political factors contribute to inequities in patient care. Combined didactic and experiential learning may be more effective to teach medical students how to address implicit bias and microaggression in the clinical setting.

**Methods:** Rising third year medical students at New York Medical College participated in a student-led implicit bias and microaggression training program, including experiential learning through standardized patient (SP) encounters and self-reflection via student-led debrief sessions. The SP encounters simulated instances of xenophobia and perceived language barriers in a hospital setting, in which students were expected to address microaggressions in real time utilizing the VITALS (Validate, Inquire, Take time, Assume the best, Leave opportunities, Speak up for others) framework.

**Results:** 200 students participated in the program. Survey responses on attitudes regarding implicit bias and microaggressions were collected prior to participating in the program (T1), after the VITALS video presentation (T2), and after the SP encounter and debrief sessions (T3). Students felt more likely to interrupt a microaggression from T1 to T3.

**Conclusions:** Our implicit bias training, equipped students with the tools and practice needed to interrupt microaggressions in the clinical setting.

**Practice Points:**

- Combined didactic and experiential learning may be more effective than didactics alone for teaching medical students how to address implicit bias and microaggression in the clinical setting.
- Students overestimated their comfort level to interrupt a microaggression after watching the training video alone, underscoring the importance of experiential learning.
- Students valued the opportunity to practice interrupting microaggressions in a safe space with the SPs.
- The SP encounter positively impacted students’ likelihood to interrupt a microaggression in the future.
- Students felt more comfortable interrupting a microaggression from a peer than from a person in power.

## Introduction

Implicit bias is defined as subconscious attitudes and thoughts that can influence one’s behavior toward, and treatment of, individuals from stereotyped groups.^1^ Such bias, particularly when present among healthcare providers, contributes to disparities in care, health status, and mortality for minoritized patients.^2^ Previous studies have indicated that implicit bias exists across all levels of the healthcare system, including medical school.^3^ Medical educators have the critical task of implementing anti-racism and anti-bias training in medical education, with the ultimate goal of mitigating the negative effects of these biases on patient care. The American Association of Medical Colleges (AAMC) has published competencies for diversity, equity and inclusion aimed at helping educators develop curricula across the medical education continuum, some of which specifically identify the need to mitigate bias in the healthcare system.^4^ Furthermore, the Liaison Committee on Medical Education (LCME) specifically requests information from medical schools regarding education experiences aimed at helping medical students recognize and address bias.^5^ Garnering awareness and understanding of implicit prejudice in healthcare early in medical education is an important initial step towards meeting the goal of mitigating the negative effects of biases on patient care. However, previous studies have underscored the need for equipping students with actionable skills and communication strategies to address bias.^6^ Learners may feel discomfort and assume avoidant behaviors to prevent patient care situations that may result in biased behavior.^6^ Anti-bias education in medical training should encourage learners to acknowledge internalized biases and their effects on patient care, as well as empower students with frameworks and communication tools for addressing such biases in clinical environments.

Several medical schools and medical training programs have implemented initiatives that encourage students to identify personal biases and provide strategies for addressing biases commonly encountered in clinical settings.^7–10^ Many workshops and training sessions have emphasized skill development through didactic lectures, case-based learning, and role play between learners. Implicit association tests before and after trainings are often used to gauge training efficacy, however the utility of such tests to assess likelihood of behavior change has been criticized.^11^ Rather than aiming to reduce implicit association test scores, some researchers suggest that implicit bias training could instead focus on enforcing positive communication behaviors through relevant opportunities to practice.^12^ However, few published curricula provide students with opportunities for experiential learning and skill development in a controlled clinical setting with peer observers and post-session discussion.

Standardized patient (SP) encounters are simulated clinical environments in which students can exercise patient care and communication strategies with trained professionals that accurately and repeatedly portray patients. Previous work has discussed the utility of SP encounters in enabling learners to practice skills such as gender-affirming care, inclusive language, and LGBTQ-inclusive sexual history taking, among others.^13–15^ Student survey evaluations in one such study demonstrated improvement in comfort with students’ ability to obtain a sexual history.^13^ The addition of peer-led discussion offers valuable opportunity for students to reflect upon the experience of applying skills and learn best practices from fellow learners. Researchers using peer-led debriefing found students to report increased comfort in discussing difficult topics and increased reassurance that conversations would remain confidential.^16^

Various models aim to provide guidance on approaching clinical situations in which the medical provider is the recipient of discriminatory remarks from patients and/or family members.^17,18^ Researchers at UCLA developed an educational tool called VITALS (Validate, Inquire, Take time, Assume the best, Leave opportunities, Speak up for others) as a framework for confronting biased statements and microaggressions that may be encountered in health care settings.^8^ Their training consisted of a workshop with didactic presentation and interactive discussion-based exercises, utilizing pre- and post-surveys to assess changes in knowledge and attitudes about microaggression intervention. In comparing pre- and post-workshop survey responses, they found that participants experienced increased comfort in broaching difficult conversations and higher likelihood of challenging microaggressions.

The goal of our curriculum was to expand upon the UCLA team’s work and develop an implicit bias training for third year medical students (MS3) at New York Medical College School of Medicine (NYMC SOM) that incorporates both traditional didactic lectures and experiential learning through SP encounters, in order to teach students how to address microaggressions in the clinical setting. To our knowledge, our study is the first to provide medical students with an opportunity to address implicit bias in the clinical setting through SP encounters followed by peer-led debrief sessions.

## Materials and Methods

All rising MS3 students in the NYMC SOM Class of 2024 participated in this mandatory educational experience, which was part of the Transition to Clerkship (TTC) program at the end of the 2021-2022 academic year. This study was approved by the New York Medical College Institutional Review Board, and our protocol #15028 was found to be exempt. Students provided consent for participation in the survey study on the first page of each Qualtrics form and the data was anonymized.

### Student Facilitator Development

A year prior to the delivery of the program, a group of eight students were selected to participate as fellows in the Transformative Educational Leadership Program (TELP), a 2-month leadership development course. This program focused on topics including leading change, self-awareness and emotional intelligence, crucial conversations, and team dynamics. The summer curriculum, occurring 2-3 times per week, varied in style between lectures from professors, flipped-classroom sessions, and group discussions. Following the summer lecture series, the team met throughout the year to develop and implement implicit bias training for rising MS3 peers. TELP students collaborated with faculty from the Office of Undergraduate Medical Education, the Office of Diversity and Inclusion, and the Clinical Skills Center to write the SP encounter scripts, design and compile pre- and post-session surveys, and prepare session debrief guides.

### Workshop Surveys

To measure the impact of our curriculum, we administered surveys at three distinct time points: prior to participating in the program (T1), after the VITALS video presentation (T2), and after the SP encounter and debrief sessions (T3) **(Appendices A-C)**. QR codes for accessing the surveys were provided at the clinical skills center before and after watching the video and after the SP encounter and debrief session.

Surveys were adapted from the UCLA VITALS study to better capture demographic information and qualitative data on the interventions deployed.^8^ Survey responses were collected on a 5-point Likert scale and administered using Qualtrics, a third party web-based survey tool for secure data collection. Each survey underwent pre-testing by ten different individuals of similar age range and educational level as our medical students. Student responses were de-identified, as students generated a non-identifiable code to allow us to link pre-session and post-session survey responses but not identify individuals to their surveys. We excluded surveys without adequate responses and surveys completed outside of the acceptable date range for the intervention (e.g. pre-intervention surveys submitted after the intervention). We analyzed the deidentified survey data using IBM SPSS Statistics software Version 28.0. We grouped together survey responses of “Agree” and “Strongly agree” in the analysis and in the data presented in the tables; such responses will be henceforth referred to as “Agreed.” Similarly, we grouped together responses of “Disagree” and “Strongly disagree,” which will henceforth collectively be referred to as “Disagreed.”

Implicit Bias Training Workshop **(Figure 1)**

**Figure 1.**
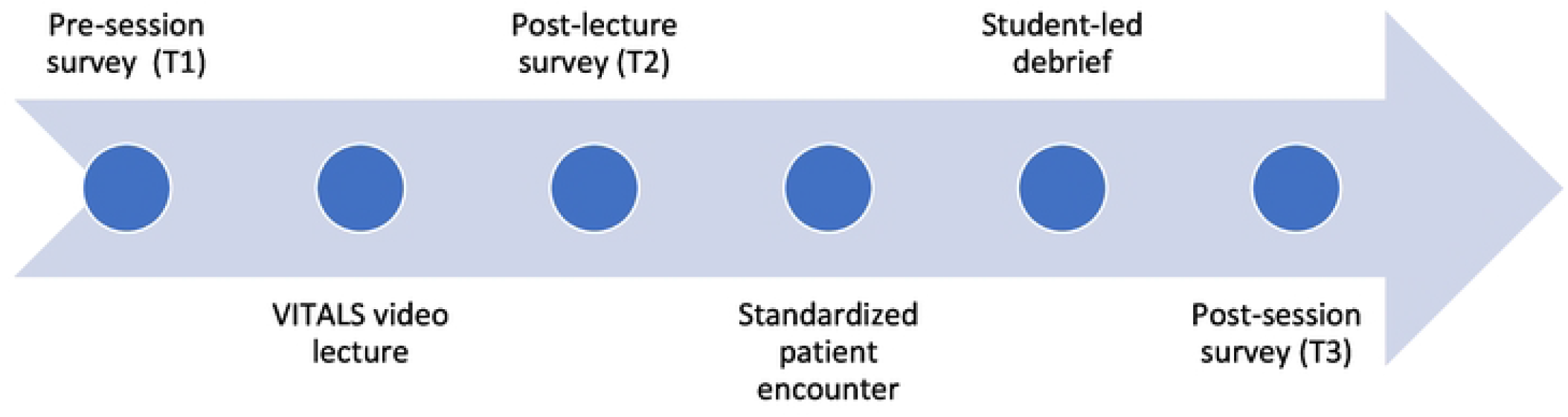
Educational Program Design.

### Pre-session Survey

We provided all MS3s with the pre-video survey via a link in the learning management system approximately 48 hours prior to the workshop.

### VITALS Video

Students were required to watch a 12-minute video lecture describing the VITALS framework **(Appendix D)** and complete a follow up post-video survey.

### Orientation

Prior to the SP encounters, students participated in a 15-minute orientation regarding the objectives and agenda of the workshop, which included ground rules to ensure safety of discussion, opportunities to discuss prior experiences with microaggressions, and a review of the VITALS framework. Students who had not yet watched the video or completed the surveys were given the opportunity to do so. Students were paired and assigned to either the participant or observer role for each of the two cases.

### SP Recruitment and Training

SPs at NYMC undergo an interview process and six months of onboarding training to obtain certification. Onboarding includes workshops on bias, assessment standard setting, history gathering and portrayal, feedback, and interpersonal communication skills. SPs are continuously monitored through data trends, review of encounters, and assessment of interrater reliability, and also participate in an annual recertification course. We developed SP scripts in collaboration with the Director of the Clinical Skills and Simulation Center (CSSC), as well as the Director of the SP Program, to ensure that formatting and encounter structure are consistent with those from SP training. The SPs for our study were selected based on availability and were provided scripts in advance to provide adequate time to review and seek clarifications as needed.

### SP Encounters and Debrief Sessions

We held SP encounters and debrief sessions at the CSSC located on campus. Each session included two rounds of encounter simulations with 14 rooms operating simultaneously. Seven rooms started with Case #1 **(Appendix E)** regarding xenophobia and perceived language barriers, and the other seven rooms started with Case #2 **(Appendix F)** regarding foreign-trained clinicians and their medical competency. Each room had two students: one student participating in the 15-minute encounter and the other student observing. After the first encounter, students participated in a 20-minute group debrief session before returning to their simulation rooms, switching roles, and participating in a second 15-minute encounter utilizing the second case the pair had not seen. The workshop concluded with a 40-minute group debrief session.

## Results

Two hundred rising MS3s at NYMC were included in this study. They completed a pre-workshop survey which assessed initial attitudes on implicit bias and collected demographic information **(Table 1)**. Approximately 50% of participants were between 25 and 26 years old, with 54.6% of students identifying as Caucasian, 30% Asian, 6.6% African American, and 8.1% other. The sample was equally divided between males and females, with 7.2% identifying with gender and sexual minorities. Regarding language, 70.4% of respondents shared that they were exposed to a language other than English in their childhood with 49.7% of those reporting always or mostly speaking another language at home, and about half of participants identified that they are proficient in a language other than English. The largest religion represented was Christianity at 29%, followed by Judaism at 21.5%. Hinduism, Islam, Agnostic, Atheism, and other religions collectively represented 15% of respondents and another 34% did not specify. In terms of medical specialty of interest, 30% of participants identified interest in a primary care specialty, whereas over 50% were interested in pursuit of specialty care, and 15% did not specify.

**Table 1.**
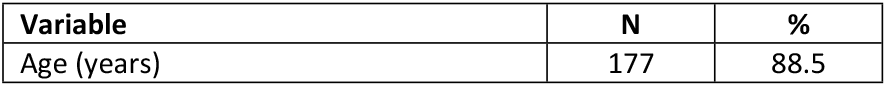
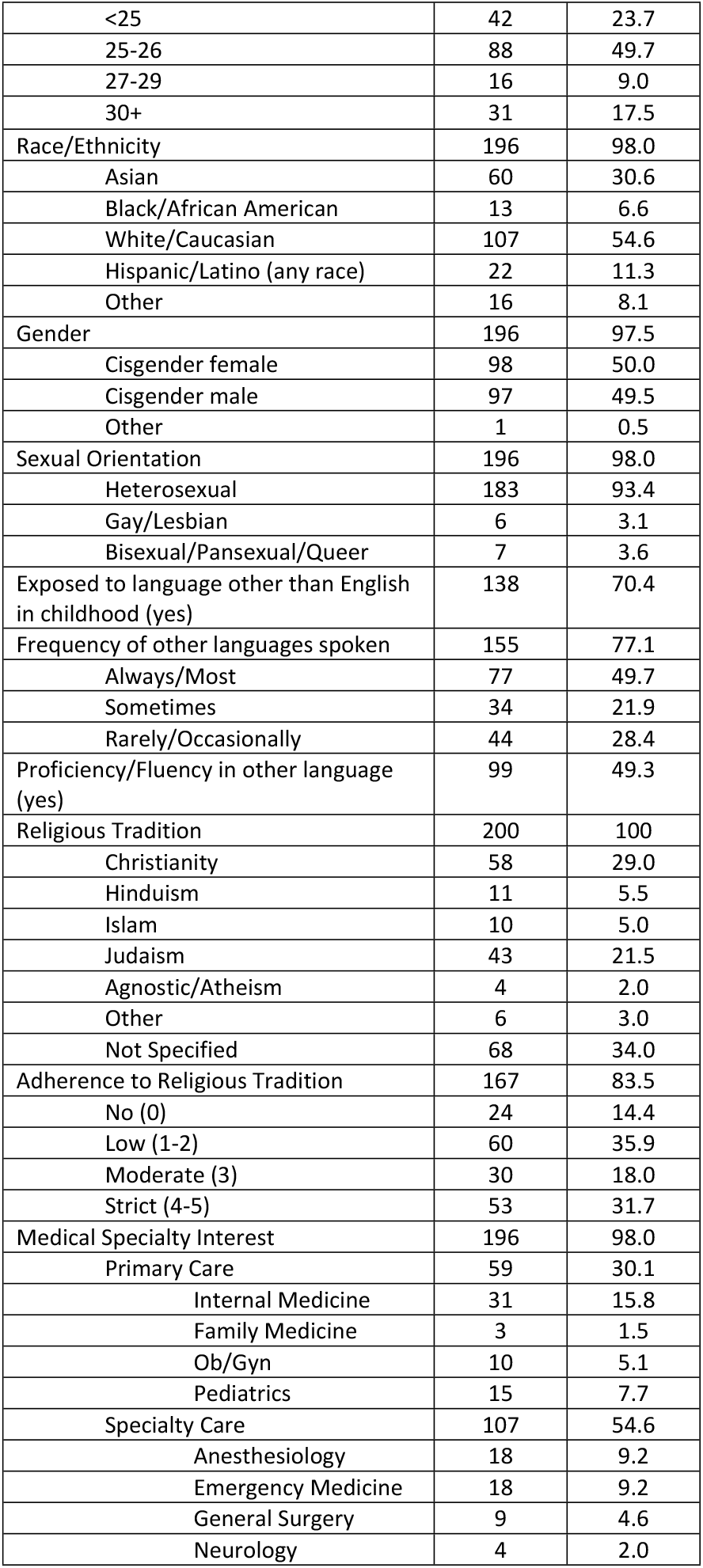
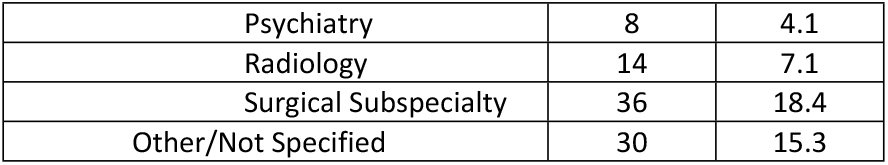
Demographic Characteristics of Study Sample (n=200).

| Variable | N | % |
| --- | --- | --- |
| Age (years) | 177 | 88.5 |
| <25 | 42 | 23.7 |
| 25-26 | 88 | 49.7 |
| 27-29 | 16 | 9.0 |
| 30+ | 31 | 17.5 |
| Race/Ethnicity | 196 | 98.0 |
| Asian | 60 | 30.6 |
| Black/African American | 13 | 6.6 |
| White/Caucasian | 107 | 54.6 |
| Hispanic/Latino (any race) | 22 | 11.3 |
| Other | 16 | 8.1 |
| Gender | 196 | 97.5 |
| Cisgender female | 98 | 50.0 |
| Cisgender male | 97 | 49.5 |
| Other | 1 | 0.5 |
| Sexual Orientation | 196 | 98.0 |
| Heterosexual | 183 | 93.4 |
| Gay/Lesbian | 6 | 3.1 |
| Bisexual/Pansexual/Queer | 7 | 3.6 |
| Exposed to language other than English in childhood (yes) | 138 | 70.4 |
| Frequency of other languages spoken | 155 | 77.1 |
| Always/Most | 77 | 49.7 |
| Sometimes | 34 | 21.9 |
| Rarely/Occasionally | 44 | 28.4 |
| Proficiency/Fluency in other language (yes) | 99 | 49.3 |
| Religious Tradition | 200 | 100 |
| Christianity | 58 | 29.0 |
| Hinduism | 11 | 5.5 |
| Islam | 10 | 5.0 |
| Judaism | 43 | 21.5 |
| Agnostic/Atheism | 4 | 2.0 |
| Other | 6 | 3.0 |
| Not Specified | 68 | 34.0 |
| Adherence to Religious Tradition | 167 | 83.5 |
| No (0) | 24 | 14.4 |
| Low (1-2) | 60 | 35.9 |
| Moderate (3) | 30 | 18.0 |
| Strict (4-5) | 53 | 31.7 |
| Medical Specialty Interest | 196 | 98.0 |
| Primary Care | 59 | 30.1 |
| Internal Medicine | 31 | 15.8 |
| Family Medicine | 3 | 1.5 |
| Ob/Gyn | 10 | 5.1 |
| Pediatrics | 15 | 7.7 |
| Specialty Care | 107 | 54.6 |
| Anesthesiology | 18 | 9.2 |
| Emergency Medicine | 18 | 9.2 |
| General Surgery | 9 | 4.6 |
| Neurology | 4 | 2.0 |
| Psychiatry | 8 | 4.1 |
| Radiology | 14 | 7.1 |
| Surgical Subspecialty | 36 | 18.4 |
| Other/Not Specified | 30 | 15.3 |

Survey responses on attitudes regarding implicit bias and microaggressions prior to the training (T1), after the VITALS video presentation (T2), and after the in-person SP encounter and debrief (T3) were collected **(Table 2)**. Response rate for T1 was 100%, for T2 was 89.5%, and T3 was 99%. At T1, 58% of respondents reported that they would challenge someone who engages in a microaggression. When further inquiring about level of comfort interrupting microaggression depending upon the setting, students were least likely (27% Agreed) to challenge a microaggression if it were to come from an individual in a position of power, and most likely (75% Agreed) to challenge a microaggression if it were made by a peer in a social setting. At T2, these reported percentages increased, with 68.2% of students reporting that they are likely to interrupt a microaggression, 83.8% indicating that they would be comfortable interrupting a microaggression made by a peer in a social setting, and 48% indicating that they would be comfortable interrupting a microaggression involving a power dynamic. At T3, 73.2% of students (increased from 58% at baseline) indicated that they would be likely to challenge someone to interrupt a microaggression. Additionally, 90.9% of students felt comfortable broaching this conversation with peers, and 40.9% felt comfortable broaching this conversation with those in power in a professional setting.

**Table 2.**
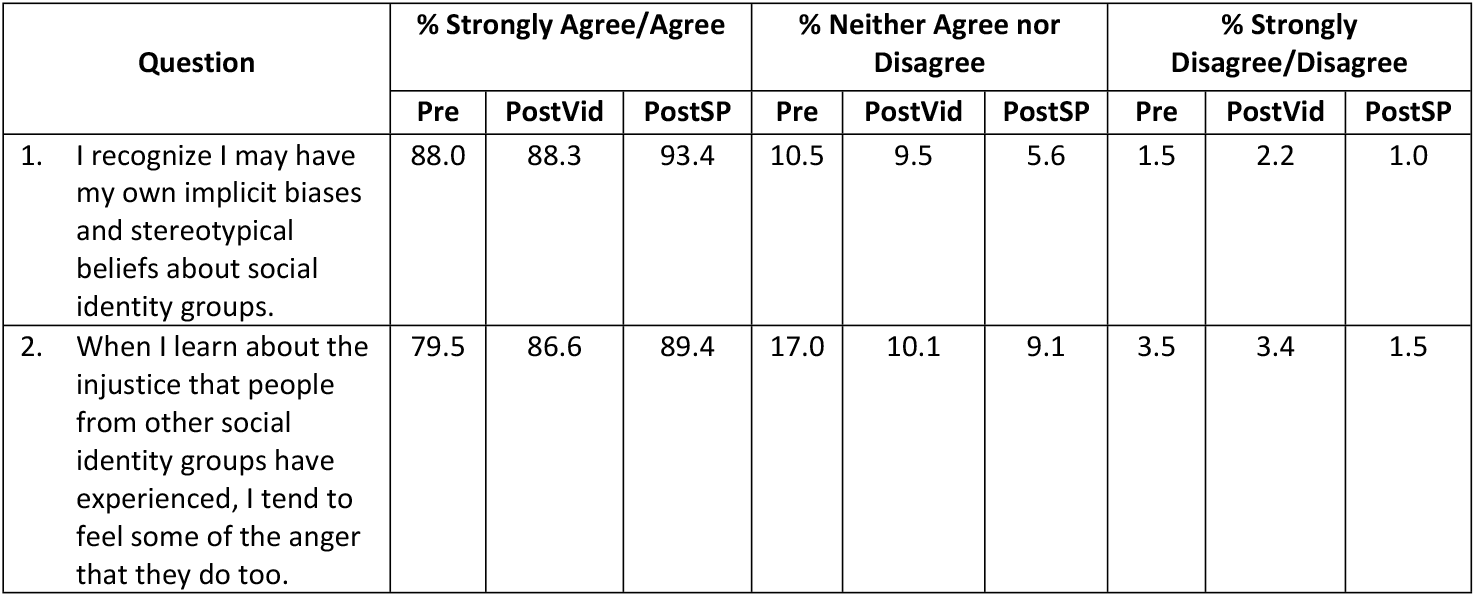
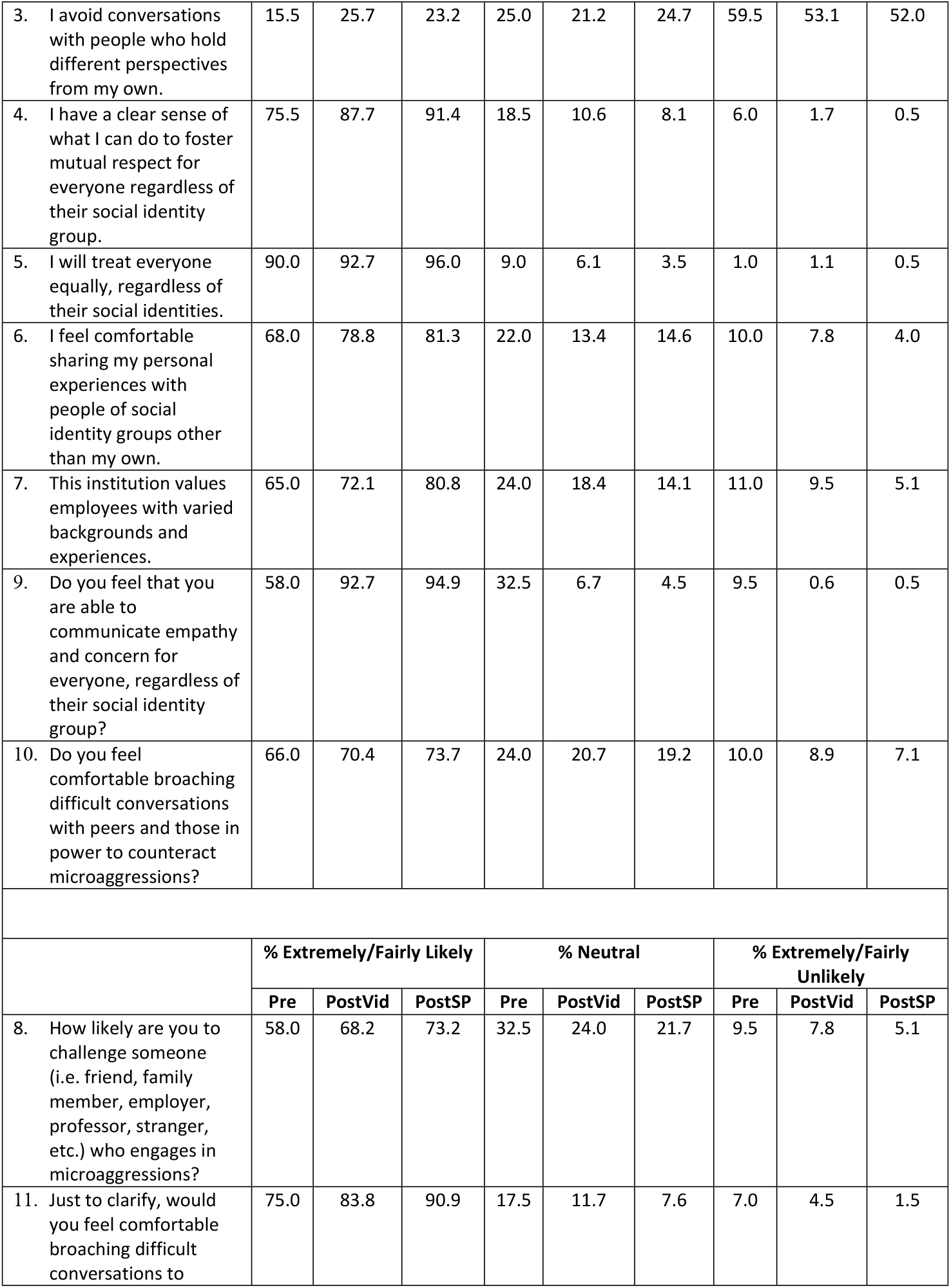
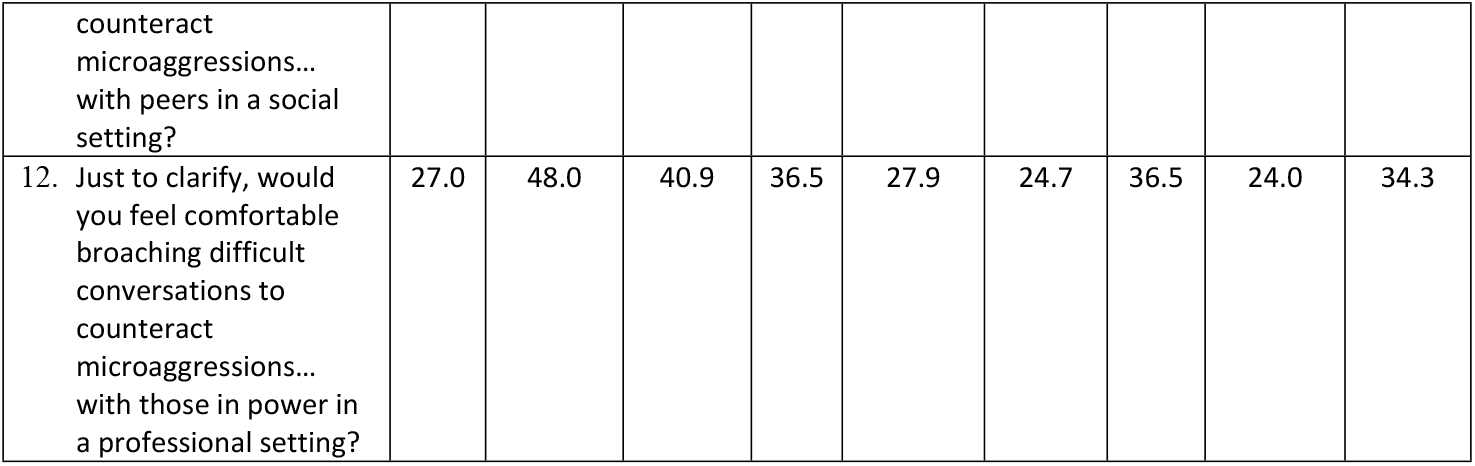
Percent Agreement with Survey Items at Pre-Survey ‘Pre’ (N=200), Post-VITALS Video ‘PostVid’ (N=179), Post-SP ‘PostSP’(N=198).

## Discussion

Our results indicate that the SP encounters positively impacted students’ likelihood to confront microaggressions. In general, nearly three quarters of the MS3 class reported that they felt more likely to interrupt a microaggression after the SP encounter than at baseline. Students reported increased comfort in their own ability to use the skills taught to interrupt a microaggression after the SP session than the video alone, except in situations where they were confronting a person in power. However, from narrative data collected during the debrief sessions, many students expressed that they experienced difficulty addressing microaggressions in the SP encounters, despite being prepared and knowing the objectives of the session. Across all debrief discussions, students reflected on how they overestimated their sense of comfort and preparedness prior to the encounters. Despite our expectation that participants would feel increasingly more comfortable with enhanced exposure to implicit bias training through simulated encounters in a safe environment, these findings highlight the importance of our multimodal approach to learning and may better reflect true participant response.

The positive change in participant attitudes from T1 to T2 aligns with previous studies showing the impact of traditional didactic lectures on student learning.^8^ As a whole, baseline pre-intervention agreement in our study was higher than survey responses in previous studies, which may be due to prior implicit bias training and/or the diversity of our student body. These increased baseline values may also impact the degree of change in agreement across the study.

Interestingly, we saw an improvement in self-reported responses in all survey question responses except for one question (Q12). After participating in the SP encounters, students responded that their level of comfort decreased with regards to situations that involve a power differential in a professional setting (27% felt comfortable at T1, 48% at T2, and 40.9% at T3). These results suggest that despite the baseline perception that participants would be comfortable interrupting microaggressions in a professional setting involving people in power, after experiencing the simulated encounter without a power dynamic at play, participants recognized potential discomfort that they may not have previously recognized. This discomfort may be attributed to the concern that speaking up towards someone in a position of power may negatively impact the student’s grade or clinical evaluation.

Previous literature focusing on the healthcare system shows pervasive adverse effects of implicit bias and systemic racism in the US healthcare system.^4,5^ Existing literature describes efforts to mitigate bias in the medical profession with education at the medical school level. To our knowledge, however, this is the first program to create an immersive experience where medical students were tasked with confronting a bias in real time with live SPs.^8-11^ Our data shows an improvement in student preparedness and comfort in confronting microaggressions in the clinical setting. The simulated clinical encounter allowed students to assess their own confidence in addressing microaggressions. Our data demonstrated a respondent shift bias highlighting the discrepancy between student perceived comfort with addressing microaggressions after a standard lecture, as compared to their actual level of comfort during a simulated encounter, underscoring the need for simulated encounters in medical education focusing on implicit bias management.

### Limitations

Since our primary outcomes were based on surveys that evaluated self-reported change in attitudes, we relied on students’ self-awareness and did not have an external measure of behavior change. Quantitative survey data collected at multiple time points are susceptible to shift bias. This may explain the overconfidence in comfort and ability at T2 and its deflation at T3. Electronic survey data can also have erroneous response selection, respondents submitting an answer choice they had not intended. We also do not have data to assess impact of the intervention beyond the immediate short term. Furthermore, our SP encounters did not include a grading rubric or checklist, which could have provided opportunity for formalized student and SP feedback after the encounter. This determination was partly at the request of the SPs who did not feel like they had adequate expertise to provide such feedback. Lastly, the design of our workshop, due to logistical constraints, was such that one student observed an encounter before participating in their own encounter may have impacted the experience and behavior of the student who engaged in the encounter secondarily.

### Future Directions

Given the importance of continued training in diversity, equity, and inclusion, we aim to expand this training to engage students in more challenging scenarios where the implicit bias or microaggression is directed at the student, or where the act is committed by an attending physician or supervisor. Introducing a personal component and power dynamic would help to further prepare students for difficult situations that they may encounter, and, as conveyed by students, would be more challenging than a microaggression alone.

Future directions also include providing additional implicit bias training specifically for the SPs to enable them to provide more structured formal feedback to students after the simulated encounters. By training the SPs, students would be able to receive real time feedback on the interaction.

Additionally, we plan to conduct focus groups with students to assess the impact of the curriculum on their clinical experiences and actions. Specifically, we would like to find out if they experienced or witnessed a microaggression and whether they addressed the microaggression in real time. Further expansion of this program to include training sessions more frequently throughout the school year may promote enhanced longitudinal education and greater commitment to core themes of the training.

## Data Availability

Data cannot be shared publicly because of the means of collection hosted on institutional Qualtrics servers. Extracted data are available by contacting Rachel Thommen after receiving approval from the NYMC IRB.

## Acknowledgements

We want to acknowledge the Standardized Patients (SPs) of New York Medical College for bringing our simulation curriculum to life.

## Appendices

A. Pre-Survey (T1)
B. Post-VITALS Video Survey (T2)
C. Post-SP Encounter and Debrief Survey (T3)
D. VITALS 12-minute Video Presentation
E. Case #1 Script
F. Case #2 Script

